# Kidney function and long-term treatment with lithium salts for bipolar disorder Determinants of mGFR and accuracy of kidney microcysts detection in the diagnosis of CKD

**DOI:** 10.1101/2021.04.11.21255136

**Authors:** Nahid Tabibzadeh, Anne-Laure Faucon, Emmanuelle Vidal-Petiot, Fidéline Serrano, Lisa Males, Pedro Fernandez, Antoine Khalil, François Rouzet, Coralie Tardivon, Nicolas Mazer, Caroline Dubertret, Marine Delavest, Emeline Marlinge, Bruno Etain, Frank Bellivier, François Vrtovsnik, Martin Flamant

## Abstract

**Background and objectives:** Kidney damage during early stages of lithium treatment in bipolar disorder is still hypothetical. We aimed at identifying the determinants of a decreased measured glomerular filtration rate (mGFR) and the accuracy of kidney MRI imaging in its detection.

**Design, setting, participants and measurements:** Cross sectional cohort study. Systematic evaluation including mGFR of 217 consecutive adult patients treated with lithium carbonate. Kidney MRI imaging in 99 patients with half-Fourier turbo spinecho and Single-shot with long echo time sequences.

**Results:** Median age was 51 [27-62] years, and median lithium treatment duration was 5 [2-14] years. 52% of patients had a stage 2 CKD. In multivariable analysis, the determinants of a lower mGFR were a longer lithium treatment duration (β −0.8 [-1; - 0.6] ml/min/1.73m^2^ GFR decrease for each year of treatment), a higher age (β −0.4 [-0.6; −0.3] ml/min/1.73m^2^ for each year of age, p<0.001), albuminuria (β −3.97 [-6.6;-1.3], p=0.003), hypertension (β −6.85 [-12.6;-1.1], p=0.02) and hypothyroidism (β −7.1 [-11.7;-2.5], p=0.003). Serum lithium concentration was not associated with mGFR. Patients with mGFR<60 ml/min/1.73m^2^ (22%) had a higher prevalence and number of renal microcysts. Renal MRI displayed renal microcyst(s) in 51% of patients, detected as early as one year after lithium treatment initiation. mGFR and lithium treatment duration were strongly correlated in patients with microcyst(s) (r=-0.64, p<0.001), but not in patients with no microcysts (r=-0.24, p=0.09). The presence of microcysts was associated with the detection of an mGFR <45 ml/min/1.73m^2^ (AUC 0.893, p<0.001, sensitivity 80%, specificity 81% for a cut-off value of 5 microcysts).

**Conclusions:** Besides usual risk factors of GFR decrease, lithium treatment duration strongly impacted mGFR independently of age, especially in patients with microcysts. Hypothyroidism was also negatively associated with kidney function in these patients. MRI might help detect early lithium-induced kidney damage and inform preventive strategies.

## Introduction

Lithium salts are the main prophylactic treatment of bipolar disorder, which is characterized by potentially life-threatening manic and/or depressive episodes. They have proven efficient in the prevention and treatment of acute episodes as well as in the prevention of suicidal risk^1–4^. However, this efficacy is counterbalanced by a narrow therapeutic range that can lead to potentially harmful overdose, and by long-term adverse events^5^. Amongst them, long-term lithium treatment might lead to polyuria and polydipsia, related to impaired urine concentrating ability, and to chronic kidney disease usually with a tubulo-interstitial presentation. The latter is characterized by mild or absent proteinuria (often < 1g/24h) and cortical and/or medullary small cysts, which supposedly appear at late stages^6^.

Epidemiological data suggest that chronic use of lithium may lead to impaired glomerular filtration rate (GFR). Indeed, in a retrospective study including 2500 patients on lithium therapy, Shine *et al*. showed an increased risk of stage 3 chronic kidney disease (CKD) defined by creatinine-based estimated GFR (eGFR)^7^. However, in a meta-analysis gathering 385 studies, McKnight *et al*. showed that eGFR was only 6.2 ml/min/1.73m^2^ lower in patients treated with lithium salts than in control populations, with an overall small risk of renal failure^8^. Another study showed that among 145 patients treated with lithium for more than 15 years, only 21% displayed a measured GFR (mGFR) below the −2 SD cut-off for age-adapted GFR^9^. It is thus still unclear whether lithium treatment at therapeutic concentrations is harmful in itself or if the GFR decrease in this population is related to age or to other comorbidities such as metabolic syndrome or hypertension as suggested by the cohort study by Clos *et al*. ^10^. If present, the magnitude of the effect of lithium treatment is also unknown.

The aim of our study was to describe clinical, biological and imaging characteristics of a cohort of lithium-treated patients focusing on renal function and radiological kidney features, and to evaluate the correlates of GFR, including the relationship between kidney function and lithium treatment, using a gold standard method for GFR measurement.

## Methods

### Study design and population

From March 2015 to December 2020, 230 consecutive adult patients were referred by psychiatrists to the Department of Renal Physiology for their first visit with nephrologists. Among these patients, we excluded 13 patients who had discontinued lithium treatment, leaving 217 patients who were referred for a systematic check-up. Eligible patients were ≥ 18 years of age at inclusion, with various durations of lithium treatment, and had neither started dialysis nor underwent kidney transplantation. The study was performed according to the Declaration of Helsinki. The study was approved by the local ethics committee from APHP.Nord (Institutional Review Board CER-2021-74). All patients provided written informed consent before inclusion in the study cohort.

### Data collection and measurements

During a 5-hour in-person visit, a large set of clinical and laboratory data were collected, including past medical history, dose and duration of lithium treatment, serum lithium concentration, and current treatment with other psychotropic drugs. Obesity was defined as body mass index (BMI) > 30 kg/m^2^. Diabetes was defined as fasting glycemia > 7 mmol/L or antidiabetic drug treatment. Hypothyroidism was either defined as a thyroid-stimulating hormone (TSH) level > 4.1 mUI/L (above the upper normal limit) or a thyroid hormone therapy. Blood pressure (BP) was the average of three measurements in resting conditions. Hypertension was defined as BP ≥140/90 mmHg or the use of antihypertensive drugs. Patients were instructed to fast (not to eat or drink) from 8 p.m. the day before the admission. Patients were asked to collect 24-hour urine the day before admission. Indications to discard first morning void on the first day, and then to collect all urine until the first void the next morning were given by a trained nurse and detailed in a written information document. Fasting blood and urine samples were collected. GFR was measured by urinary clearance of ^51^Cr-EDTA or ^99^Tc-DTPA (GE Healthcare, Velizy, France and Curium, Saclay, France respectively) ^11^, determined as the average of 7 consecutive 30-minutes urinary clearance periods, indexed to the standard body surface area of 1.73m^2^ as previously described^12^. Plasma lithium concentration was the last available measurement during the last 6 months. Creatinine was measured using an Isotopic Dilution Mass Spectroscopy-standardized enzymatic method.

### Kidney MRI

A renal magnetic resonance imaging (MRI) was proposed to all the patients, and was finally performed in 99 patients due to the remaining patients’ refusal (mainly for claustrophobia or unavailability). MRI was performed with a pre-specified protocol using the following sequences: T2 SSFSE (T2 weighted sequence single-shot fast spin-echo), also called half-Fourier single-shot turbo spin-echo (or HASTE for Siemens), and SSFSE TE long (Single-shot fast spin echo with long echo time). T2 SSFSE is an ultrafast MRI technique with a short acquisition time less susceptible to motion respiratory artifact than other techniques such as echo T2-weighted imaging. SSFSE TE long is a highly T2 weighted sequence allowing an optimal contrast between microcysts and renal parenchyma. This MRI protocol has proven useful in depicting structures containing static fluids including cystic lesions, characterized as T2 hyperintense round areas (figure 3) ^13^.

Renal microcysts were defined as small (1–2 mm) round cystic lesions and quantified in both kidneys. As inter and intra-observer reproducibility was poor when counting the total number of microcysts, a semi-quantitative approach was used, and MRI findings were classified as follows: 1/ no microcysts, 2/ 1 to 5 microcysts, 3/ 6 to 10 microcysts, 4/ 11 to 20 microcysts, 5/ 21 to 50 microcysts, 6/ 51 to 100 microcysts, 7/ > 101 microcysts. Images were blindly reviewed and analyzed by 2 senior radiologists and one senior nephrologist. With this semi-quantitative scoring, reproducibility was 100%.

### Statistical analyses

Categorical variables were described as frequencies and percentages, and continuous variables as median [25^th^-75^th^ percentiles]. Baseline patients’ characteristics were compared according to mGFR classes (>90, 60-90, <60 mL/min/1.73m^2^) using Kruskal–Wallis test and Chi-square tests, for quantitative and qualitative variables, respectively. Correlations between mGFR, age and lithium treatment duration (log-transformed) were assessed by Spearman tests. Determinants of mGFR were assessed using multivariable linear regression models. Covariates were selected *a priori* based on potential confounding and included: age, gender, body mass index (< vs ≥ 30kg/m^2^), diabetes, hypertension, hypothyroidism, treatment with other psychotropic drugs, lithium treatment duration in years (log-transformed), lithium galenic formulation (extended or immediate-release formulation), serum lithium concentration and 24-hour urinary albumin/creatinine ratio (log-transformed). Interactions between treatment duration and age, gender, and BMI were tested. Normality of the distribution of residuals and homoscedasticity were verified. Multiple imputations using the chained equation method (R package *mice*, n=50 imputed datasets, m=20 iterations) were performed for missing data.

In order to specifically analyze the association between mGFR and renal microcysts, a Spearman correlation was performed between mGFR and lithium treatment duration in patients with microcysts and those without microcysts. A linear regression model was built to explain mGFR according to treatment duration, with an interaction term between treatment duration and renal microcysts to assess the difference in mGFR slopes between patients with and without microcysts. The effect of microcysts on slopes was estimated and a Wald test was performed on the interaction term to compare the two groups.

With an mGFR < 45 ml/min/1.73m^2^ considered as the state of disease, sensitivity (Se) and specificity (Sp) were calculated for each microcyst cut-off score. A Receiver Operating Characteristic (ROC) Curve was plotted in order to measure the Area Under the Curve (AUC) and the Youden Index allowed identifying the most accurate cut-off value for renal microcysts.

A two-sided p-value < 0.05 was considered statistically significant. Statistical analyses were conducted using in GraphPad Prism 9.0 and R 3.4 software.

## Results

### Characteristics of the population

Table 1 summarizes patients’ characteristics. Median age was 51 [27-62] years, and 62% were female. Median BMI was 25.9 [22.9-28.8] kg/m^2^. Median lithium treatment duration was 5 [2-14] years and median dose was 800 [575-1000] mg/day. The extended-release form was administered in a majority of patients (63%). Median plasma lithium concentration was 0.72 [0.56-0.87] mmol/l. Median mGFR was 78 [63-91] ml/min/1.73m^2^ with a majority of stage 2 CKD (52%). Microalbuminuria was absent (< 3 mg/mmol) in a large majority (83%) of patients and only 2 patients had a macroalbuminuria (which remained below 0.6 g/24h). Seven (3%) patients had diabetes mellitus, 26% had hypertension and 36% had hypothyroidism.

**Table 1.** Characteristics of the patients in the whole cohort and according to measured GFR.

| Variables, n<br>available data | total<br>n=217 | mGFR > 90<br>ml/min/1.73m<sup>2</sup><br>n=57 | 60≤mGFR<90<br>n=113 | mGFR<60<br>ml/min/1.73m<sup>2</sup><br>n=47 | p-value |
| --- | --- | --- | --- | --- | --- |
| Age, years n=217 | 51 [37-62] | 38 [31-48.5] | 51 [28-61.5] | 62 [54.5-69] | <0.001 |
| Male patients,<br>n=217 | 83 (38.2%) | 33 (57.9%) | 38 (33.3%) | 12 (10.5%) | <0.001 |
| Body mass index,<br>kg/m<sup>2</sup>, n=217 | 25.9 [22.9-28.8] | 25.5 [23.1-29.3] | 25.6 [22.5-29] | 26.8 [23.6-28.8] | 0.7 |
| Diabetes mellitus,<br>n=217 | 7 (3.2) | 1 (1.8%) | 4 (3.5%) | 2 (4.3%) | 0.7 |
| Hypertension,<br>n=217 | 56 (25.8) | 6 (10.5%) | 27 (23.7%) | 23 (48.9%) | <0.001 |
| Hypothyroidism,<br>n=217 | 78 (35.9) | 12 (21.1%) | 41 (36%) | 23 (53.2) | 0.01 |
| Other psychotropic<br>drugs, n=216 | 152 (70.1%) | 35 (61.4%) | 83 (73.5%) | 34 (72.3%) | 0.3 |
| Lithium treatment<br>duration, years,<br>n=217 | 5 [2-14] | 2 (1-5) | 5 (2-12) | 16 [9.5-30] | <0.001 |
| Extended-release<br>formulation, n=214 | 138 (63.6%) | 41 (71.9%) | 74 (64.9%) | 23 [48.9%] | 0.04 |
| Daily dose, mg/day<br>n=216 | 800 [575-1000] | 1000 [788-1200] | 800 [750-1000] | 500 [400-750] | <0.001 |
| Serum lithium,<br>mmol/l, n=139 | 0.72 [0.56-0.87] | 0.72 [0.53-0.9] | 0.75 [0.6-0.9] | 0.61 [0.5-0.84] | 0.4 |
| mGFR,<br>ml/min/1.73m<sup>2</sup><br>n=217 | 78 [62.8-90.5] | 97.3 [94.1-102.9] | 77.3 [71.5-83.4] | 46.6 [41-53.7] | <0.001 |
| ACR, mg/mmol<br>creatinine, n=214 | 1.4 [0.9-2.2] | 0.9 [0.8-1.9] | 1.4 [1.0-2.0] | 2.0 [1.4-4.3] | <0.001 |
| PCR, mg/mmol<br>creatinine, n=216 | 14.1 [10.1-20.2] | 11.4 [8.7-15.3] | 14.0 [10.1-19.7] | 19.6 [15.6-33.2] | <0.001 |
| renal MRI findings | n=99 | n=28 | n=48 | n=23 |  |
| no microcysts | 49 (49.4%) | 20 (71.4%) | 26 (54.2%) | 3 (13%) | <0.001 |
| 1-10 microcysts | 30 (30.3%) | 7 (25%) | 15 (31.3%) | 8 (34.8%) | <0.001 |
| > 10 microcysts | 20 (20.2%) | 1 (3.6%) | 7 (14.6%) | 12 (52.2%) | <0.001 |

Categorical and continuous data are expressed in n (%) and in median [quartile 1- quartile 3], respectively. Baseline patients' characteristics were compared across mGFR level using the chi-2 test for categorical variables and Kruskal-Wallis Rank Sum Test for quantitative variables. mGFR: measured glomerular filtration rate, ACR: urinary albumin to creatinine ratio, PCR: urinary protein to creatinine ratio, MRI: magnetic resonance imaging.

Compared with patients with mGFR higher than 60ml/min/1.73m^2^, patients in the lowest mGFR group (< 60 ml/min/1.73m^2^) were significantly older, with a higher prevalence of hypertension, and were more frequently female. In this population, lithium treatment duration was significantly longer, and daily dose was significantly lower with a lower rate of extended-release lithium preparation. Of note, there was no statistical difference according to GFR groups for serum lithium levels. Even though levels were low in all groups, they also displayed a higher albuminuria and higher proteinuria (table 1).

The prevalence of CKD was higher in patients with a longer treatment duration (figure 1). In patients treated for less than one year, only 2 patients had an mGFR < 60 ml/min/1.73m^2^. Among patients treated for more than 25 years, none had a normal mGFR (>90 ml/min/1.73m^2^).

**Figure 1.**
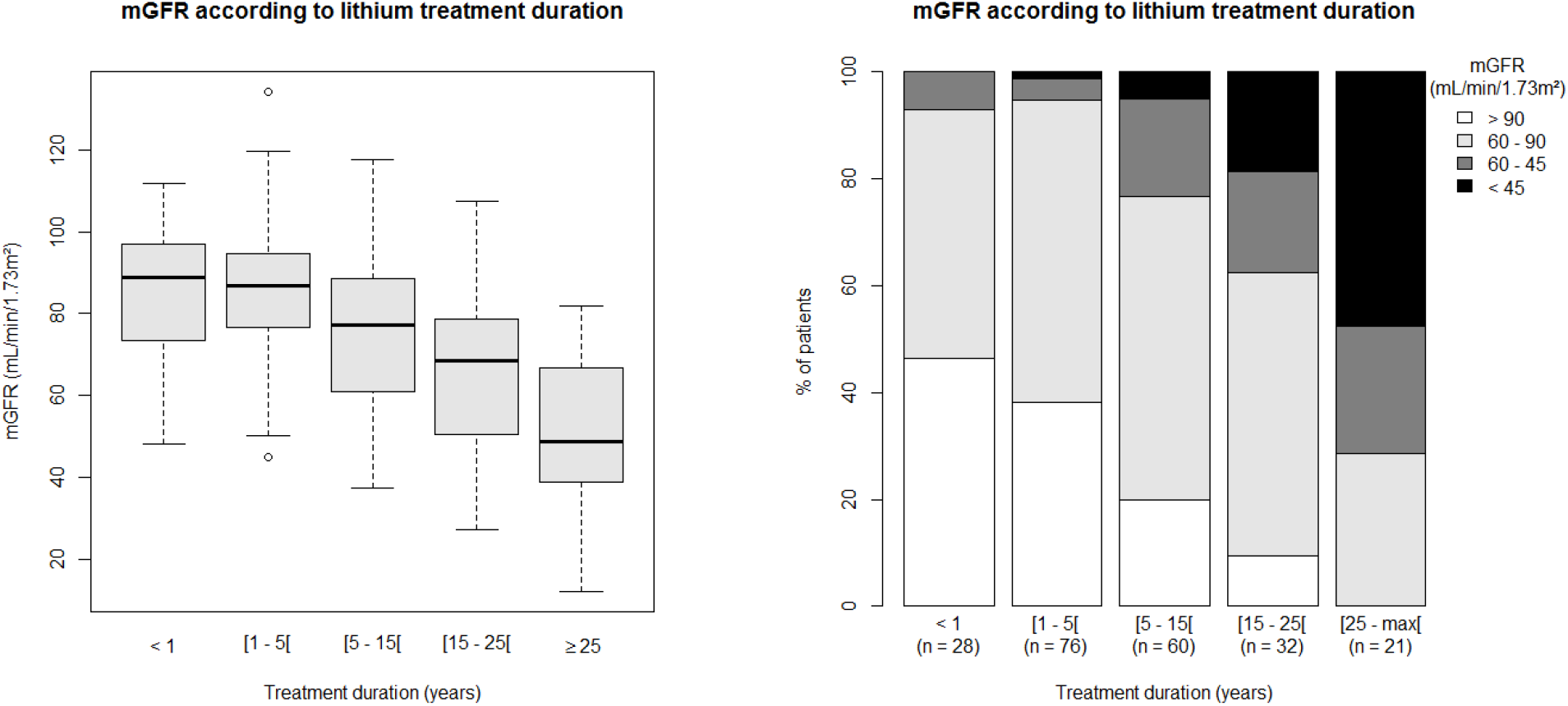
mGFR according to lithium treatment duration. Left panel: median and range of mGFR according to lithium treatment duration. Right panel: stacked bars representing mGFR classes according to lithium treatment duration. mGFR: measured glomerular filtration rate.

### Determinants of mGFR

Lithium treatment duration (log-transformed) and age were inversely correlated with mGFR (r=-0.51, p<0.001 and r=-0.54, p<0.001, respectively). In multivariable analysis, lithium treatment duration and age were also independently and negatively associated with mGFR, with a stronger association with lithium treatment duration (β-coefficient: −0.8 [-1; −0.6] ml/min/1.73m^2^ GFR decrease for each additional year of treatment) compared to age (β-coefficient: −0.4 [-0.6; −0.3] ml/min/1.73m^2^ for each additional year of age, p<0.001). Association between treatment duration and mGFR did not depend on age or other covariates (non-significant p-values for interaction tests) (figure 2). Albuminuria, hypothyroidism and hypertension were also independently and negatively associated with mGFR (β-coefficients respectively - 3.97 [-6.6;-1.3], p=0.003, −7.1 [-11.7;-2.5], p=0.003, −6.85 [-12.6;-1.1], p=0.02). Diabetes was associated with a higher mGFR, but was diagnosed in only 7 patients including one with hyperfiltration. The other tested covariables - including serum lithium concentration, the use of extended-release form of lithium carbonate and treatment with other psychotropic agents - were not independently associated with mGFR.

**Figure 2.**
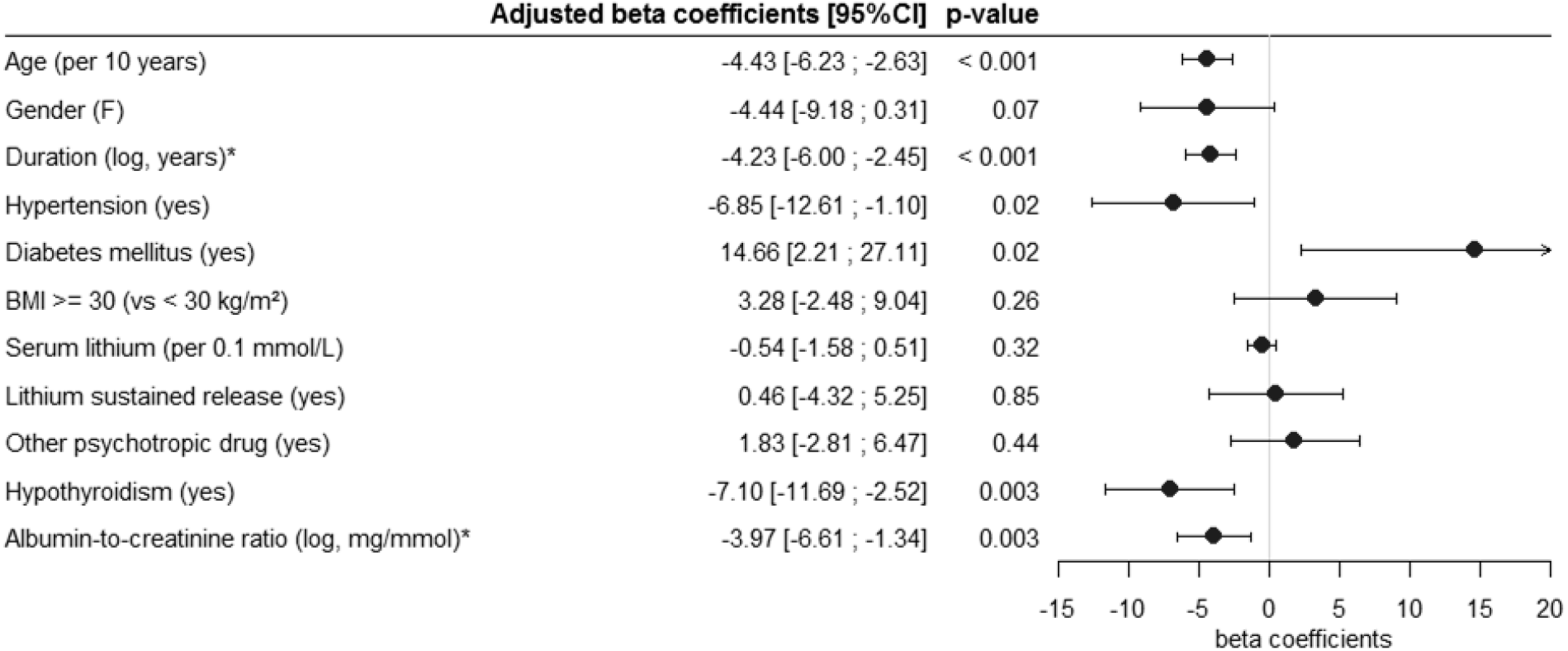
Multivariable analysis of the determinants of mGFR. Determinants were assessed using multivariable linear regression model, with multiple imputations for missing data. All the variables included in the model are represented on the forest plot. F: female, BMI: body mass index, mGFR: measured glomerular filtration rate. * The indicated coefficients correspond to a 2.72-fold [=exp(1)] increase in lithium treatment duration or in albumin-to-creatinin ratio.

### Accuracy of renal microcysts for the diagnosis of lithium-related nephrotoxicity

Among patients who underwent renal MRI 51% displayed renal microcyst(s), appearing as T2 hyperintense round lesions uniformly and symmetrically distributed throughout the medulla as well as the cortex of normal-sized kidneys. Isolated Bosniak type 1 cysts were also unfrequently observed. Microcysts were present in patients as soon as one year after lithium treatment initiation. Patients in the lowest mGFR group had a higher prevalence of renal microcysts. When comparing patients with and without microcysts, mGFR and lithium treatment duration were strongly correlated in patients with microcyst(s) (r=-0.64, p<0.001), whereas no such correlation was found in patients with no microcysts (r=-0.24, p=0.09). This was confirmed by the linear regression of mGFR according to lithium treatment duration in patients with microcysts (slope −1.15, 95% CI [-1.44; −0.86]) and with no microcysts (slope −0.30, 95% CI [-1.03; 0.43]), with a statistically significant difference between the slopes of the two groups (Wald test p=0.02) (figure 3).

Using receiver operating characteristic (ROC) analysis, we determined the renal microcyst quantification threshold associated with the detection of an mGFR below 45 ml/min/1.73m^2^ (figure 3). The AUC was 0.893 (p<0.001). At a cut-off value of 5 microcysts, the Youden Index was maximal (0.61), with a Se and a Sp respectively of 80% and 81%.

**Figure 3.**
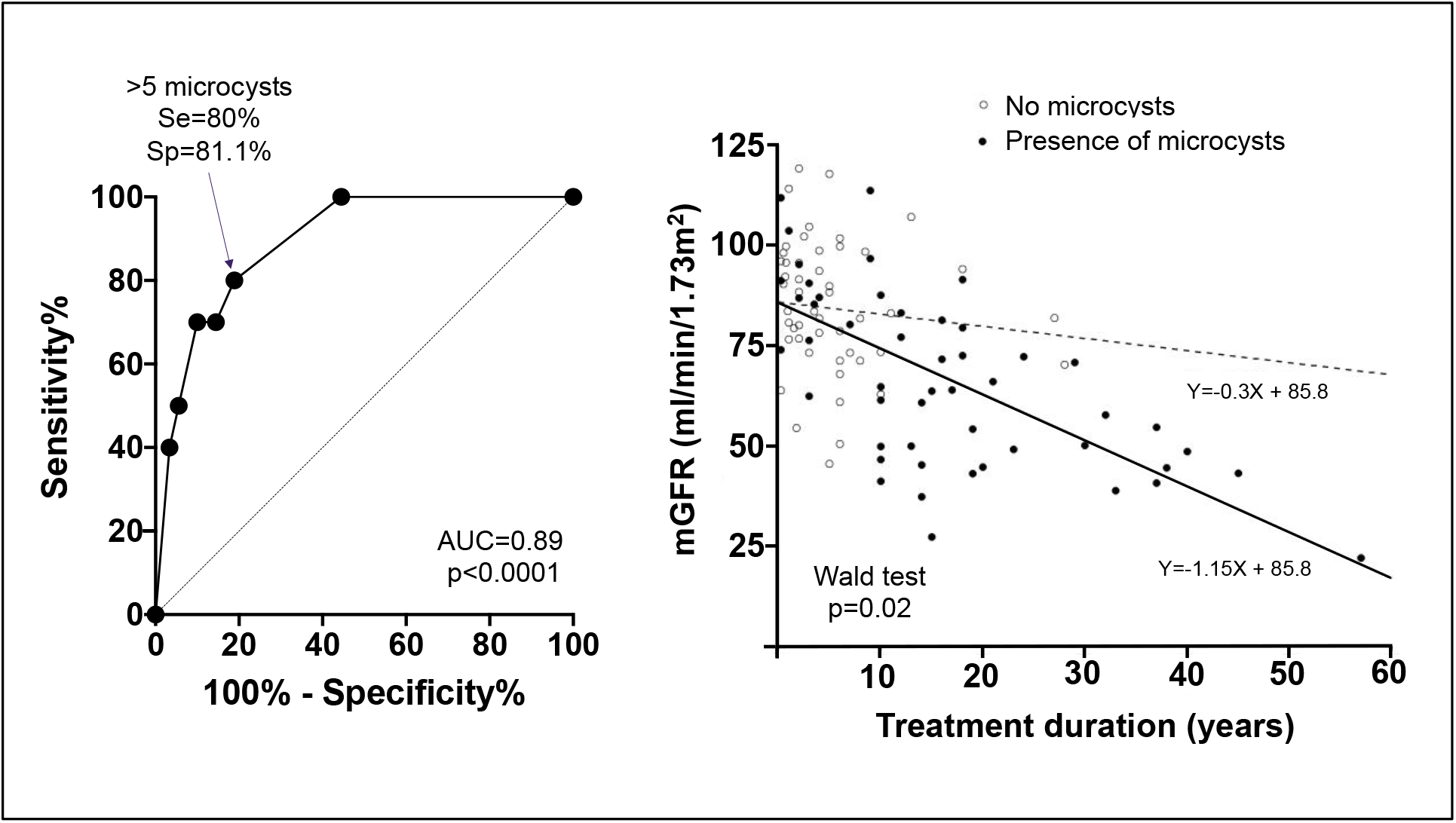
Accuracy of renal microcysts in the diagnosis of lithium-related chronic kidney disease. **Left panel:** ROC Curve showing the sensitivity and 1 - specificity of renal microcysts quantification for the diagnosis of mGFR < 45 ml/min/1.73m^2^. The cut-off value of 5 microcysts had the highest Youden Index, with a sensitivity (Se) of 80% and a specificity (Sp) of 81.1%. **Right panel**: GFR values according to treatment duration and regression lines in patients with (filled circles and black line) and without (open circles and dashed line) microcysts. A Wald test was performed to test the difference in slopes between the two groups.

## Discussion

Our study showed a strong relationship between chronic lithium use and GFR decline, independently of other potential confounding factors. The specific effect of lithium use on GFR decrease was determined as a 0.8 ml/min/1.73m^2^ mean decrease in mGFR per year of exposition. Other determinants of a lower mGFR were higher age, higher albuminuria, hypertension and hypothyroidism.

There are conflicting results regarding renal effects of lithium salts^5^. Several reports suggest that even when CKD is present, it is not clinically significant as no increase in the risk of ESKD has been demonstrated ^14–16^. In their meta-analysis, McKnight *et al*. demonstrated a slight eGFR reduction (mean −6.2 ml/min/1.73m^2^) in patients treated with lithium compared to control groups^8^. The main limitation of these studies is the short follow-up duration and the short median treatment duration. In a large retrospective study of laboratory analyses from more than 2500 patients, Shine *et al*. observed an increased risk of CKD stage 3 in lithium-treated patients compared to control populations ^7^. In the same line, a French retrospective study showed that 39% of patients aged 20-39 years, and up to 85% in patients aged 70 years or older displayed decreased GFR (defined as an estimated GFR < 60 ml/min/1.73m^2^)^17^. Accordingly, a large Swedish cohort study showed that 1.2% of lithium-treated patients had a serum creatinine level > 150 µmol/l ^18^. As controversial as kidney impairment during chronic lithium use may be, the general view is that lithium treatment induces a slowly progressive tubulo-interstitial nephropathy, that cannot be easily detected in longitudinal studies ^19^.

In this perspective, evolution towards end-stage kidney disease (ESKD) is also poorly defined. A retrospective study among patients on dialysis showed that 0.22% of these patients had a diagnosis of lithium-induced nephropathy^20^. In the Swedish cohort, the prevalence of ESKD in lithium-treated patients was 0.5% - more than 6 times the estimated prevalence in the general population^18^ - after a mean treatment duration of 23 years.

Conflicting results have also been reported in experimental studies regarding lithium effect on kidney ^21^. As few studies have experimented long-term exposure to lithium, a satisfying experimental model of lithium-induced nephropathy with renal insufficiency is still lacking in order to clarify the renal impact of chronic lithium use.

Our study allowed establishing a clear relationship between lithium use and GFR decrease. Indeed, in this population, the strongest determinant of mGFR was lithium treatment duration. The effect of lithium treatment duration was twice that of age, though a cumulative effect of age and treatment duration might potentiate renal lesions secondary to lithium chronic use on an underpinning senescent renal tissue. Of note, a majority of the patients were treated with the extended-release formulation of lithium carbonate, which is supposed to decrease the risk of kidney disease by limiting the risk of acute rises of serum lithium levels ^22–24^. Noteworthy, a higher serum lithium level was not associated with lower mGFR but the high proportion of missing data (36% of missing serum lithium values) might have limited the statistical power of the study. Our results are in line with the study by Bendz *et al*. showing no difference in terms of plasma lithium levels between patients with CKD and ESKD and those with normal renal function ^9^. In contrast, Shine *et al*. suggested a higher risk of adverse renal outcomes with higher serum lithium levels, but the analysis was not adjusted on baseline eGFR ^7^. In any case, a causal relationship between serum lithium level and mGFR should be interpreted with caution in observational studies, as lower mGFR is also responsible of lower excretion rate of lithium and thus higher serum lithium levels ^21^. Consistently, in our study, the daily dose of lithium was higher in patients with higher mGFR, which is not surprising as serum lithium level depends on lithium glomerular filtration which guides drug dose adaptation. Moreover, a single plasma lithium measurement is probably not representative of the long-term cumulative exposure.

Age was also an independent determinant of mGFR. Age is a well-demonstrated determinant of GFR decline. However, GFR decline over time might reflect kidney senescence rather than pathological processes^25^. The combined effect of age and lithium treatment is likely superior to this physiological GFR decrease over time.

Lithium administration is associated with metabolic disorders including weight gain and thyroid disorders, the pathophysiology of which are not fully understood^26^. Obesity is a risk factor for CKD and ESKD ^27^. Other factors influencing BMI such as thyroid disorders^28^ might also promote kidney disease^29^. We thus tested whether obesity or hypothyroidism might determine mGFR in these patients. Our analyses ruled out the role of obesity but found a strong association between hypothyroidism and lower mGFR. As previously reported, hypothyroidism was frequent in our study population (36%). The literature suggests a higher prevalence of hypothyroidism in female patients treated with lithium salts, with no reported effect of treatment duration on that risk. However, to the best of our knowledge, the association between kidney impairment and hypothyroidism in patients treated with lithium had not yet been described. Thyroid hormones exert both direct and indirect effects on renal functions, including cardiovascular effects affecting renal hemodynamics ^28^ potentially explaining the link between hypothyroidism and renal risk reported in previous epidemiological data ^30^. However, our observational study does not establish causality between hypothyroidism and a lower mGFR, and the hypothesis of an association due to a common pathway of susceptibility to kidney and thyroid toxicities induced by chronic lithium treatment cannot be excluded.

The majority of patients (83%) displayed mild CKD with no albuminuria, and only 2 patients had a macroalbuminuria (< 0.6 g/24h), suggesting that glomerular disease did not participate in the kidney disease and that CKD was mostly not related to other underlying nephropathies. Yet, albuminuria was associated with a lower mGFR in agreement with current knowledge ^31^. The prevalence of diabetes was also very low (3%) in our cohort, and one patient displayed glomerular hyperfiltration, thus inducing a bias in the association between mGFR and diabetes. Conversely, hypertension was observed in 23% of patients and was an independent determinant of mGFR. It has been shown that CKD both contributes to the development of hypertension, and might result from hypertension^32^. However, the prevalence of hypertension was far lower in our population compared to previously reported CKD populations ^33^, and even less than the reported prevalence in the general population in France ^34^. Consequently, our results suggest that though hypertension is a relevant comorbidity contributing to the decrease in mGFR in patients treated with lithium salts, it is probably not a feature of lithium-induced nephropathy.

Renal cysts have been reported during long-term lithium use. However, the majority of published data involve small case series or case reports of patients with numerous microcysts and overt chronic kidney disease ^35–37^. It is thus not yet clearly established whether renal microcysts are a specific and early feature of kidney impairment of chronic lithium use and if they might be used as a diagnostic tool. In our cohort we found that even a relatively small number of microcysts was associated with a decrease in mGFR, with acceptable sensitivity and specificity to detect an mGFR < 45 ml/min/1.73m^2^. The optimal cut-off value was 5 microcysts. Of major importance, while there was a strong negative correlation between mGFR and lithium treatment duration in patients with at least one microcyst, no such association was seen in patients without microcysts, in favor of a strong relationship between mGFR decrease and the presence of microcysts. The presence of microcysts might thus help inform improved strategies such as decreasing lithium exposure and preventing other comorbidities and nephrotoxic agents. These results must however be interpreted with caution due to the sample size (n=99). Further investigation is also needed to establish if they reflect the degree of irreversibility, as the potential benefit of treatment discontinuation is still poorly known ^9,38^ and shall be weighed against the established suicidal risk in this setting ^39^.

Our study displays some limitations. Regarding lithium treatment, serum lithium level was not measured the same day as renal evaluation. However, our patients were stable, and underwent regular follow-up by psychiatrists. Second, neither information regarding cumulative lithium dose nor episodes of overdose were available. We were thus not able to investigate whether nephrotoxic effect of lithium was related to acute serum lithium level rises, or to a cumulative effect of low dose exposition^23^. Of note, previous data suggest that renal impairment during chronic lithium use is not related to cumulative lithium dose ^9^. Finally, the cross-sectional design of the study prevents us from analyzing the predictive value of lithium treatment duration on mGFR decline. Also, we cannot exclude that lithium was initiated in some patients with an already altered GFR, but it can be noted that only 2 out of 42 patients treated with lithium for less than one year had an mGFR below 60 ml/min/1.73m^2^.

In conclusion, our study confirmed the independent effect of lithium exposure on kidney function. This effect combined with that of age, hypothyroidism, hypertension and microalbuminuria on mGFR demonstrates that a close monitoring is necessary including blood pressure measurements, GFR estimation, albuminuria and thyroid hormone levels. MRI might also be considered as a useful tool in order to detect microcysts even during early stages of treatment. The association of a lower GFR with microcysts and hypothyroidism questions the issue of a differential individual genetic or acquired susceptibility to lithium treatment. These data might help inform improved strategies to prevent irreversible kidney damage during chronic lithium use.

## Data Availability

The data are available upon reasonable request to the corresponding author.

## Acknowledgements

The authors wish to acknowledge the patients who accepted to enter the study, the nursing and medical staff who took care of the patients, and the technical staff who performed the imaging and the radioisotope measurements.

## Bibliography

1. Geddes JR, Burgess S, Hawton K, Jamison K, Goodwin GM. Long-term lithium therapy for bipolar disorder: systematic review and meta-analysis of randomized controlled trials. Am J Psychiatry. 2004;161(2):217–222. doi:10.1176/appi.ajp.161.2.217

2. BALANCE investigators and collaborators, Geddes JR, Goodwin GM,et al. Lithium plus valproate combination therapy versus monotherapy for relapse prevention in bipolar I disorder (BALANCE): a randomised open-label trial. Lancet Lond Engl. 2010;375(9712):385–395. doi:10.1016/S0140-6736(09)61828-6

3. Cipriani A, Hawton K, Stockton S, Geddes JR. Lithium in the prevention of suicide in mood disorders: updated systematic review and meta-analysis. BMJ. 2013;346:f3646.

4. Miura T, Noma H, Furukawa TA, et al. Comparative efficacy and tolerability of pharmacological treatments in the maintenance treatment of bipolar disorder: a systematic review and network meta-analysis. Lancet Psychiatry. 2014;1(5):351–359. doi:10.1016/S2215-0366(14)70314-1

5. Tabibzadeh N, Vrtovsnik F, Serrano F, Vidal-Petiot E, Flamant M. [Chronic metabolic and renal disorders related to lithium salts treatment]. Rev Med Interne. 2019;40(9): 599-608. doi:10.1016/j.revmed.2019.01.006

6. Grünfeld J-P, Rossier BC. Lithium nephrotoxicity revisited. Nat Rev Nephrol. 2009;5(5):270–276. doi:10.1038/nrneph.2009.43

7. Shine B, McKnight RF, Leaver L, Geddes JR. Long-term effects of lithium on renal, thyroid, and parathyroid function: a retrospective analysis of laboratory data. Lancet Lond Engl. 2015;386(9992):461–468. doi:10.1016/S0140-6736(14)61842-0

8. McKnight RF, Adida M, Budge K, Stockton S, Goodwin GM, Geddes JR. Lithium toxicity profile: a systematic review and meta-analysis. Lancet Lond Engl. 2012;379(9817):721–728. doi:10.1016/S0140-6736(11)61516-X

9. Bendz H, Aurell M, Balldin J, Mathé AA, Sjödin I. Kidney damage in long-term lithium patients: a cross-sectional study of patients with 15 years or more on lithium. Nephrol Dial Transplant Off Publ Eur Dial Transpl Assoc - Eur Ren Assoc. 1994;9(9):1250–1254.

10. Clos S, Rauchhaus P, Severn A, Cochrane L, Donnan PT. Long-term effect of lithium maintenance therapy on estimated glomerular filtration rate in patients with affective disorders: a population-based cohort study. Lancet Psychiatry. 2015;2(12):1075–1083. doi:10.1016/S2215-0366(15)00316-8

11. Vidal-Petiot E, Courbebaisse M, Livrozet M, et al. Comparison of 51Cr-EDTA and 99mTc-DTPA for glomerular filtration rate measurement. J Nephrol. Published online March 4, 2021. doi:10.1007/s40620-020-00932-9

12. Froissart M, Rossert J, Jacquot C, Paillard M, Houillier P. Predictive Performance of the Modification of Diet in Renal Disease and Cockcroft-Gault Equations for Estimating Renal Function. J Am Soc Nephrol. 2005;16(3):763–773. doi:10.1681/ASN.2004070549

13. Nakayama Y, Yamashita Y, Matsuno Y, et al. Fast breath-hold T2-weighted MRI of the kidney by means of half-Fourier single-shot turbo spin echo: comparison with high resolution turbo spin echo sequence. J Comput Assist Tomogr. 2001;25(1):55–60. doi:10.1097/00004728-200101000-00010

14. Kessing LV, Gerds TA, Feldt-Rasmussen B, Andersen PK, Licht RW. Use of Lithium and Anticonvulsants and the Rate of Chronic Kidney Disease: A Nationwide Population-Based Study. JAMA Psychiatry. 2015;72(12):1182. doi:10.1001/jamapsychiatry.2015.1834

15. Melick EJM van, Meinders AE, Hoffman TO, Egberts TCG. Renal effects of long-term lithium therapy in the elderly: a cross-sectional study. Int J Geriatr Psychiatry. 2008;23(7):685-692. doi:https://doi.org/10.1002/gps.1961

16. Aprahamian I, Santos FS, Santos B dos, et al. Long-Term, Low-Dose Lithium Treatment Does Not Impair Renal Function in the Elderly: A 2-Year Randomized, Placebo-Controlled Trial Followed by Single-Blind Extension. J Clin Psychiatry. 2014;75(7):672–678. doi:10.4088/JCP.13m08741

17. Bassilios N, Martel P, Godard V, et al. Monitoring of glomerular filtration rate in lithium-treated outpatients--an ambulatory laboratory database surveillance. Nephrol Dial Transplant Off Publ Eur Dial Transpl Assoc - Eur Ren Assoc. 2008;23(2):562–565. doi:10.1093/ndt/gfm567

18. Bendz H, Schön S, Attman P-O, Aurell M. Renal failure occurs in chronic lithium treatment but is uncommon. Kidney Int. 2010;77(3):219–224. doi:10.1038/ki.2009.433

19. Davis J, Desmond M, Berk M. Lithium and nephrotoxicity: a literature review of approaches to clinical management and risk stratification. BMC Nephrol. 2018;19(1):305. doi:10.1186/s12882-018-1101-4

20. Presne C, Fakhouri F, Noël L-H, et al. Lithium-induced nephropathy: Rate of progression and prognostic factors. Kidney Int. 2003;64(2):585–592. doi:10.1046/j.1523-1755.2003.00096.x

21. Alsady M, Baumgarten R, Deen PMT, de Groot T. Lithium in the Kidney: Friend and Foe? J Am Soc Nephrol JASN. 2016;27(6):1587–1595. doi:10.1681/ASN.2015080907

22. Castro VM, Roberson AM, McCoy TH, et al. Stratifying Risk for Renal Insufficiency Among Lithium-Treated Patients: An Electronic Health Record Study. Neuropsychopharmacol Off Publ Am Coll Neuropsychopharmacol. 2016;41(4):1138–1143. doi:10.1038/npp.2015.254

23. Carter L, Zolezzi M, Lewczyk A. An updated review of the optimal lithium dosage regimen for renal protection. Can J Psychiatry Rev Can Psychiatr. 2013;58(10):595–600. doi:10.1177/070674371305801009

24. Turan T, Eşel E, Tokgöz B, et al. Effects of short-and long-term lithium treatment on kidney functioning in patients with bipolar mood disorder. Prog Neuropsychopharmacol Biol Psychiatry. 2002;26(3):561–565. doi:10.1016/s0278-5846(01)00308-6

25. Delanaye P, Jager KJ, Bökenkamp A, et al. CKD: A Call for an Age-Adapted Definition. J Am Soc Nephrol. 2019;30(10):1785–1805. doi:10.1681/ASN.2019030238

26. Gracious BL, Meyer AE. Psychotropic-Induced Weight Gain and Potential Pharmacologic Treatment Strategies. Psychiatry Edgmont. 2005;2(1):36–42.

27. Chang AR, Zafar W, Grams ME. Kidney Function in Obesity – Challenges in Indexing and Estimation. Adv Chronic Kidney Dis. 2018;25(1):31–40. doi:10.1053/j.ackd.2017.10.007

28. Mariani LH, Berns JS. The Renal Manifestations of Thyroid Disease. J Am Soc Nephrol. 2012;23(1):22–26. doi:10.1681/ASN.2010070766

29. Brenner BM, Meyer TW, Hostetter TH. Dietary protein intake and the progressive nature of kidney disease: the role of hemodynamically mediated glomerular injury in the pathogenesis of progressive glomerular sclerosis in aging, renal ablation, and intrinsic renal disease. N Engl J Med. 1982;307(11):652–659. doi:10.1056/NEJM198209093071104

30. Rhee CM, Kalantar-Zadeh K, Streja E, et al. The relationship between thyroid function and estimated glomerular filtration rate in patients with chronic kidney disease. Nephrol Dial Transplant. 2015;30(2):282–287. doi:10.1093/ndt/gfu303

31. Levey AS, Gansevoort RT, Coresh J, et al. Change in Albuminuria and GFR as End Points for Clinical Trials in Early Stages of CKD: A Scientific Workshop Sponsored by the National Kidney Foundation in Collaboration With the US Food and Drug Administration and European Medicines Agency. Am J Kidney Dis Off J Natl Kidney Found. 2020;75(1):84–104. doi:10.1053/j.ajkd.2019.06.009

32. Ku E, Lee BJ, Wei J, Weir MR. Hypertension in CKD: Core Curriculum 2019. Am J Kidney Dis. 2019;74(1):120–131. doi:10.1053/j.ajkd.2018.12.044

33. Vidal-Petiot E, Metzger M, Faucon A-L, et al. Extracellular Fluid Volume Is an Independent Determinant of Uncontrolled and Resistant Hypertension in Chronic Kidney Disease: A NephroTest Cohort Study. J Am Heart Assoc. 2018;7(19):e010278. doi:10.1161/JAHA.118.010278

34. Perrine AL, Lecoffre C, Blacher J, Olié V. NATIONAL PREVALENCE OF HYPERTENSION, TREATMENT AND CONTROL, IN FRANCE IN 2015 AND TEMPORAL TRENDS SINCE 2006. J Hypertens. 2018;36:e227. doi:10.1097/01.hjh.0000539640.04317.12

35. Farres MT, Ronco P, Saadoun D, et al. Chronic lithium nephropathy: MR imaging for diagnosis. Radiology. 2003;229(2):570–574. doi:10.1148/radiol.2292020758

36. Golshayan D, Nseir G, Venetz J-P, Pascual M, Barbey F. MR imaging as a specific diagnostic tool for bilateral microcysts in chronic lithium nephropathy. Kidney Int. 2012;81(6):601. doi:10.1038/ki.2011.449

37. Judge PK, Winearls CG. The utility of magnetic resonance imaging in the diagnosis of chronic lithium nephropathy. QJM Int J Med. 2015;108(1):75–76. doi:10.1093/qjmed/hcu138

38. Markowitz GS, Radhakrishnan J, Kambham N, Valeri AM, Hines WH, D’agati VD. Lithium Nephrotoxicity A Progressive Combined Glomerular and Tubulointerstitial Nephropathy. J Am Soc Nephrol. 2000;11(8):1439–1448.

39. Baldessarini RJ, Tondo L, Hennen J. Effects of lithium treatment and its discontinuation on suicidal behavior in bipolar manic-depressive disorders. J Clin Psychiatry. 1999;60 Suppl 2:77-84; discussion 111-116.

